# Geographical Variation in Antimalarial Drug Resistance Marker Prevalence Across the Southern African Elimination Eight Region

**DOI:** 10.64898/2026.05.20.26353165

**Authors:** Jaishree Raman, Andrés Aranda-Díaz, Maxwell Mabona, Mukosha Chisenga, Maria Florinda João, Domingos Jandondo, Pedro Rafael Dimbu, Nomcebo Nhlengethwa, Sabelo V. Dlamini, Lydia Eloff, Stark Katokele, Davis R. Mumbengegwi, Qedusizi Nyawo, Mbavhalelo Shandukani, Sydney Mwanza, Moonga Hawela, Simone Boene, Arlindo Chidimatembue, Bernardete Rafael, Eduard Rovira-Vallbona, Brighton Mangena, Sonja B. Lauterbach, Takalani I. Makhanthisa, Hazel Gwarinda, Blaženka D Letinić, Faith De Amaral, Isobel Routledge, Beatriz Arregui-Gallego, Mishalan Moodley, Jonathan Featherstone, Power B. Tshikae, Arshad Ismail, Jose Franco Martins, Quinton Dlamini, Baltazar Candrinho, Petrina Uusiku, Ednah Baloyi, Busiku Hamainza, Alfredo Mayor, Bryan Greenhouse, Amy Wesolowski, Chadwick Sikaala, John Chimumbwa, Jennifer L. Smith

## Abstract

Global efforts to control and eliminate malaria are threatened by the emergence and spread of antimalarial drug resistance. The World Health Organization recommends surveillance of molecular markers of resistance as a complementary approach to therapeutic efficacy studies. Here, we report the first regional analysis of malaria drug resistance markers from genomic surveillance across six southern African countries spanning diverse transmission intensities and geographies. Dried blood spots, collected from rapid diagnostic test-positive individuals in Angola, Eswatini, Namibia, and Zambia in 2023, Mozambique in 2022, and South Africa between 2022 and 2024 using a standardized collection method, were analyzed using a *Plasmodium falciparum* targeted amplicon sequencing protocol.

The distribution of resistance markers was spatially heterogeneous. Markers of artemisinin partial resistance (ART-R) were rare, never detected in >1.3% of samples from any country. However, over 30% of samples from eastern Namibia and Zambia’s Western and Central Provinces carried the candidate *kelch13* P441L ART-R marker. Other ART-R markers (*kelch13* R515K, P553L, P574L, A675V) were detected at low frequencies in all countries except Mozambique. The wild type *mdr1* N86 allele, potentially associated with reduced lumefantrine susceptibility, was near fixation across all countries. The sulfadoxine-pyrimethamine (SP) resistance *dhps*-*dhfr* quintuple mutation was approaching fixation in most districts, except northern Angola, where *dhps* K540E prevalence was lower and the *crt* K76T chloroquine resistance marker more frequently detected. This pronounced spatial heterogeneity underscores the need for timely high-resolution local resistance data generation and sharing to safeguard antimalarial drug efficacy and guide malaria control and elimination strategies across the region.

## Background

The already dampening progress against malaria since 2016 (WHO 2021) was further exacerbated by the COVID-19 pandemic.^1,2^ In 2024, there were approximately 9 million more cases compared to 2023, the majority from the sub-Saharan Africa.^3^ Reasons for this stagnation and, in some cases, reversal in progress include reduced funding for malaria control, disruptions in the delivery of routine malaria case management and vector control services, the spread of invasive vector species, and reduced efficacy of core interventions as a consequence of vector behavior changes and/or genetic evolution of malaria parasites and vectors.^2,4^

Antimalarial drug-resistant *Plasmodium falciparum* has historically undermined malaria control efforts in sub-Saharan Africa. During the expansion of chloroquine resistance in the region, some countries experienced a two to three-fold increase in malaria deaths and hospital admissions.^5^ Similarly, the establishment of sulfadoxine-pyrimethamine (SP)-resistant parasites in southern Africa played a pivotal role in the 1999/2000 malaria outbreak, which resulted in South Africa reporting over 60,000 cases.^6^ Retrospective genomic analyses of African chloroquine- and SP-resistant parasites revealed a close genetic relationship between parasites that emerged along the Thai-Cambodia border and those present in Africa, confirming the spread of these parasites from South East Asia through selective genetic sweeps.^7–9^ The widespread replacement of these failing antimalarials with highly effective artemisinin-based combination therapies (ACTs) played a central role in the significant reductions in malaria burden observed between 2000 and 2015.^10^ However, the recent de novo emergence of artemisinin partial resistance (ART-R) in multiple African countries threatens the sustained efficacy of ACTs and the achievement of global targets outlined in the Global Strategy for Malaria.^4,11^ Widespread resistance to ACTs in Africa is estimated to result in millions of additional malaria cases, thousands of excess deaths, and annual economic losses ranging from $100 million to $1.1 billion.^12^ Urgent action is therefore needed to protect the sustained efficacy of ACTs while new non-artemisinin-based antimalarials are brought to market.^13^ Although the World Health Organization (WHO) recommends the routine monitoring of validated molecular markers of resistance, molecular data from southern Africa remain scarce.^14,15^ A major contributing factor has been the limited regional capacity to generate, analyze, and translate molecular data into actionable information for effective programmatic decision-making.

The Southern African Development Community’s Malaria Elimination Eight (SADC E8), a consortium of eight southern African countries (Angola, Botswana, Eswatini, Mozambique, Namibia, South Africa, Zambia, and Zimbabwe), was established to promote collaborative efforts toward malaria elimination.^16^ Strengthening malaria molecular surveillance capacity and generating relevant, timely molecular data in the E8 is critical to enhancing preparedness and identifying emerging threats to ongoing control and elimination efforts. Therefore, five E8 countries (Angola, Eswatini, Namibia, South Africa, and Zambia) together with the E8 Secretariat and technical partners, including the University of California San Francisco (UCSF), Johns Hopkins University, and the South African National Institute for Communicable Diseases (NICD), implemented a regional malaria genomic epidemiology initiative, the Genomics for Malaria in the E8 (GenE8). This initiative aimed to generate programmatically relevant genomic data across the five participating countries, promote prompt data sharing, and encourage the incorporation of genomic surveillance into routine malaria surveillance activities.

The Regional GenE8 (RegGenE8) study, embedded within the GenE8 initiative, focused on generating malaria genomic data for the two high-burden countries (Angola and Zambia) and three low-burden countries (Eswatini, Namibia, and South Africa), using harmonized sampling frameworks and centralized sequencing. Concurrently, through the GenMoz project, Mozambique generated malaria molecular data comparable to that produced through the RegGenE8 study.^17^ By integrating and reanalyzing data from these two projects, this paper provides the first regional synthesis of antimalarial drug resistance marker presence and distribution across six southern African countries, highlighting their implications for malaria control and elimination efforts.

## Methods

### Study Settings

The RegGenE8 study was conducted across five malaria-endemic southern African countries, Angola, Eswatini, Namibia, South Africa, and Zambia, while the GenMoz study was conducted across nine provinces in Mozambique (Figure 1); detailed descriptions of each country setting are provided in the respective country-level analyses. ^18–23^ Malaria transmission intensity in these six countries is heterogeneous, with a general north to south decrease in transmission intensity. Although transmission occurs year-round, peak transmission typically coincides with the hot, rainy season from December to May, with seasonality becoming more pronounced as one moves further south from the Equator. *Plasmodium falciparum* is the dominant malaria parasite across the region, with *Anopheles funestus*, *An. gambiae* s.s., and *An. arabiensis* the predominant vectors. Indoor residual spraying and long-lasting insecticidal nets are the primary vector control interventions.

**Figure 1:**
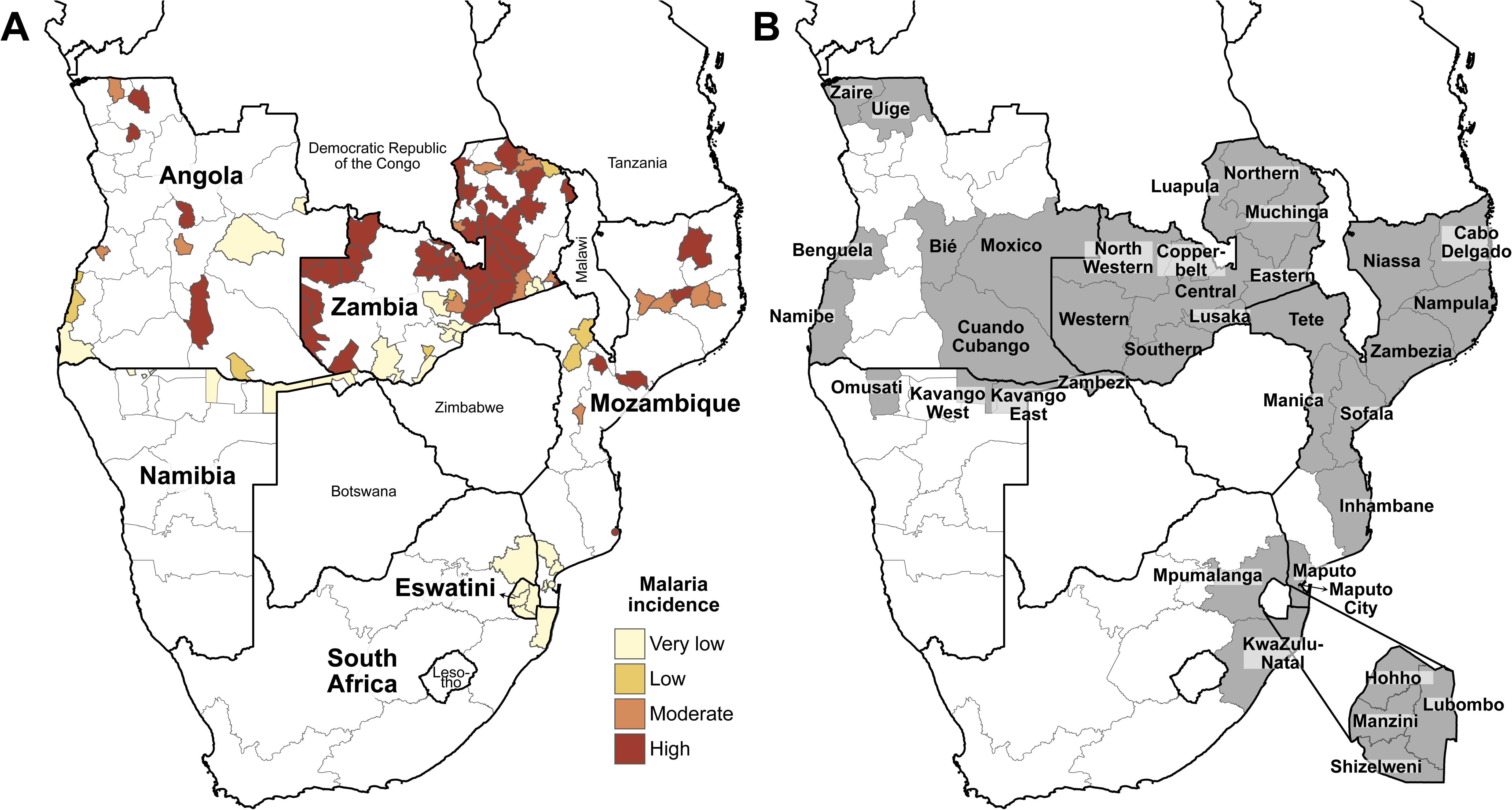
**A.** Districts from the six participating Elimination Eight countries included in this manuscript, stratified by malaria incidence according to the World Health Organization framework for malaria elimination, based on annual parasite incidence per 1,000 population: very low (<100), low (100-249), moderate (250-449), and high (≥450).^24^ Data are from 2023 for all countries except Mozambique, for which 2022 data were used, and South Africa, for which 2022-2024 data were used. **B.** Map of the southern Africa region showing the provinces or regions from which samples were collected, highlighted and labeled by country.

Rapid diagnostic tests (RDTs) and microscopy are the main diagnostic tools in the participating countries. Mozambique, South Africa and Zambia use histidine-rich protein 2 (HRP2) based *falciparum*-specific RDTs, while in Eswatini and Namibia RDTs that detect *P*. *falciparum* HRP2 and pan-*Plasmodium* lactate dehydrogenase (LDH) are recommended. Angola uses a *P*. *falciparum* HRP2 and *P*. *vivax* LDH-based RDT. Artemether-lumefantrine is a first-line ACT for uncomplicated malaria throughout the region, with artesunate-amodiaquine an alternate first-line treatment option in Angola and Mozambique. Dihydroartemisinin-piperaquine (DHAP) is also listed as a first-line treatment option in Angola. Severe malaria cases are treated with intravenous artesunate in all six countries, with intravenous quinine reserved as a second-line option in certain countries. The transmission-blocking antimalarial, single low-dose primaquine, is part of the standard treatment for uncomplicated malaria in eliminating districts of Eswatini, Namibia, and South Africa. Additional details on country-specific study settings are described in previously published country-level analyses. ^18–23^

### Study Design, Sampling, and Data Collection

The RegGenE8 study was a cross-sectional, multi-site healthcare facility-based observational study embedded within the national malaria surveillance systems of the six participating countries. The study was conducted in Angola (7 of 18 provinces), Eswatini (all regions), Namibia (all endemic regions), and Zambia (all 10 provinces) in 2023, and South Africa (2 of 3 endemic provinces) from 2022 to 2024. In Angola, logistical constraints limited sampling to a stratified subset of endemic provinces, while in South Africa, the endemic province of Limpopo did not participate in the study. Healthcare facilities were selected to capture the range of transmission settings and geographic regions within each country. The GenMoz study conducted healthcare facility-based surveys in 10 of the 11 Mozambican provinces during the 2021 and 2022 rainy seasons, as well as in two low transmission provinces during the dry season. This report only includes GenMoz data from 2022.

As sampling strategies and data collection methodologies are described in detail in the country-specific publications; a summary is provided here for completeness.^18–23^ Across all study sites, symptomatic *P. falciparum* cases confirmed by RDT or microscopy were recruited. Cases identified through passive case detection in all participating countries, as well as reactive or proactive screening in South Africa, Eswatini, and southern Zambia, were included in the study. Finger-prick dried blood spots (DBS) or used RDTs were collected from consenting individuals (see Ethical Approvals), labeled with unique barcodes linking survey and laboratory data, air-dried, and stored individually with desiccant in sealed bags until shipment for sequencing. RegGenE8 samples were shipped to either the NICD or UCSF, while GenMoz samples were transported to the Manhiça Health Research Center, Manhiça, Mozambique, or to ISGlobal in Barcelona, Spain. Demographic, clinical, and travel-history data were collected using electronic or paper-based questionnaires. Written informed consent was obtained from all participants, except in South Africa (see Ethical Approvals). Patients with severe malaria, severe malnutrition, or febrile illness attributable to non-malarial diseases were excluded from the study.

Country-specific sampling strategies and numbers of sequenced samples are summarized in Table 1. These harmonized datasets were used for downstream analysis, including mapping the prevalence and distribution of antimalarial resistance markers. Sample numbers reported here may differ slightly from those in previously published country-specific analyses, as all available RegGen8 and GenMoz sequences from the six participating countries were reanalyzed for this paper using standardized quality filters and cut-offs.

**Table 1.**
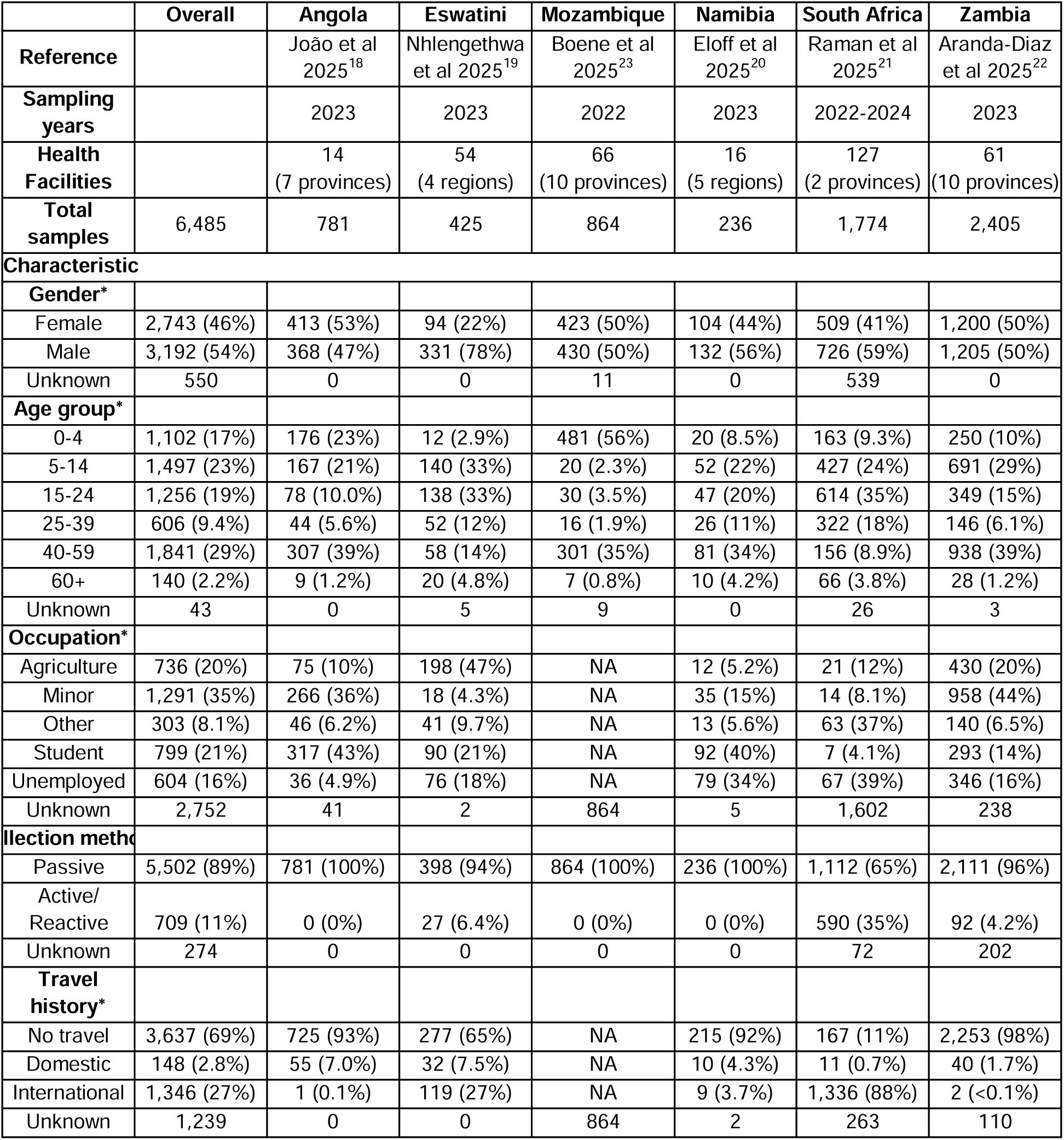
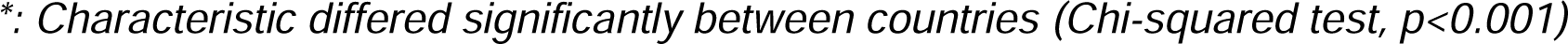
Sociodemographic characteristics of study participants in the six participating Elimination Eight countries.

|  | Overall | Angola | Eswatini | Mozambique | Namibia | South Africa | Zambia |
| --- | --- | --- | --- | --- | --- | --- | --- |
| **Reference** |  | João et al 2025<sup>18</sup> | Nhlengethwa et al 2025<sup>19</sup> | Boene et al 2025<sup>23</sup> | Eloff et al 2025<sup>20</sup> | Raman et al 2025<sup>21</sup> | Aranda-Diaz et al 2025<sup>22</sup> |
| **Sampling years** |  | 2023 | 2023 | 2022 | 2023 | 2022-2024 | 2023 |
| **Health Facilities** |  | 14<br>(7 provinces) | 54<br>(4 regions) | 66<br>(10 provinces) | 16<br>(5 regions) | 127<br>(2 provinces) | 61<br>(10 provinces) |
| **Total samples** | 6,485 | 781 | 425 | 864 | 236 | 1,774 | 2,405 |
| **Characteristic** |  |  |  |  |  |  |  |
| **Gender\*** |  |  |  |  |  |  |  |
| Female | 2,743 (46%) | 413 (53%) | 94 (22%) | 423 (50%) | 104 (44%) | 509 (41%) | 1,200 (50%) |
| Male | 3,192 (54%) | 368 (47%) | 331 (78%) | 430 (50%) | 132 (56%) | 726 (59%) | 1,205 (50%) |
| Unknown | 550 | 0 | 0 | 11 | 0 | 539 | 0 |
| **Age group\*** |  |  |  |  |  |  |  |
| 0-4 | 1,102 (17%) | 176 (23%) | 12 (2.9%) | 481 (56%) | 20 (8.5%) | 163 (9.3%) | 250 (10%) |
| 5-14 | 1,497 (23%) | 167 (21%) | 140 (33%) | 20 (2.3%) | 52 (22%) | 427 (24%) | 691 (29%) |
| 15-24 | 1,256 (19%) | 78 (10.0%) | 138 (33%) | 30 (3.5%) | 47 (20%) | 614 (35%) | 349 (15%) |
| 25-39 | 606 (9.4%) | 44 (5.6%) | 52 (12%) | 16 (1.9%) | 26 (11%) | 322 (18%) | 146 (6.1%) |
| 40-59 | 1,841 (29%) | 307 (39%) | 58 (14%) | 301 (35%) | 81 (34%) | 156 (8.9%) | 938 (39%) |
| 60+ | 140 (2.2%) | 9 (1.2%) | 20 (4.8%) | 7 (0.8%) | 10 (4.2%) | 66 (3.8%) | 28 (1.2%) |
| Unknown | 43 | 0 | 5 | 9 | 0 | 26 | 3 |
| **Occupation\*** |  |  |  |  |  |  |  |
| Agriculture | 736 (20%) | 75 (10%) | 198 (47%) | NA | 12 (5.2%) | 21 (12%) | 430 (20%) |
| Minor | 1,291 (35%) | 266 (36%) | 18 (4.3%) | NA | 35 (15%) | 14 (8.1%) | 958 (44%) |
| Other | 303 (8.1%) | 46 (6.2%) | 41 (9.7%) | NA | 13 (5.6%) | 63 (37%) | 140 (6.5%) |
| Student | 799 (21%) | 317 (43%) | 90 (21%) | NA | 92 (40%) | 7 (4.1%) | 293 (14%) |
| Unemployed | 604 (16%) | 36 (4.9%) | 76 (18%) | NA | 79 (34%) | 67 (39%) | 346 (16%) |
| Unknown | 2,752 | 41 | 2 | 864 | 5 | 1,602 | 238 |
| **Recollection method** |  |  |  |  |  |  |  |
| Passive | 5,502 (89%) | 781 (100%) | 398 (94%) | 864 (100%) | 236 (100%) | 1,112 (65%) | 2,111 (96%) |
| Active/<br>Reactive | 709 (11%) | 0 (0%) | 27 (6.4%) | 0 (0%) | 0 (0%) | 590 (35%) | 92 (4.2%) |
| Unknown | 274 | 0 | 0 | 0 | 0 | 72 | 202 |
| **Travel history\*** |  |  |  |  |  |  |  |
| No travel | 3,637 (69%) | 725 (93%) | 277 (65%) | NA | 215 (92%) | 167 (11%) | 2,253 (98%) |
| Domestic | 148 (2.8%) | 55 (7.0%) | 32 (7.5%) | NA | 10 (4.3%) | 11 (0.7%) | 40 (1.7%) |
| International | 1,346 (27%) | 1 (0.1%) | 119 (27%) | NA | 9 (3.7%) | 1,336 (88%) | 2 (<0.1%) |
| Unknown | 1,239 | 0 | 0 | 864 | 2 | 263 | 110 |

\*: Characteristic differed significantly between countries (Chi-squared test, $p < 0.001$ )

#### Sample Processing and Genomic Assays

Dried blood spot and RDT samples were processed and sequenced using previously described harmonized protocols.^18–23,25^ Genomic DNA was extracted using the Chelex-Tween 20 protocol, with parasitemia confirmed and quantified using a qPCR assay targeting the *P*. *falciparum* var gene acidic terminal sequence (varATS) in the RegGenE8 study or the 18S ribosomal RNA subunit gene in the GenMoz study.^26–28^ The extracted quantified DNA was stored at −20 °C until sequencing. Amplicon-based sequencing was performed using the MAD^4^HatTeR multiplex panel with CleanPlex reagents, using the MAD^4^HatTeR primer pools D1.1, R1.1 or R1.2, and R2.1.^25^ Sequencing was conducted on Illumina MiSeq, NextSeq or NovaSeq instruments using 150 paired-end reads.

Allele calls were obtained using the bioinformatic pipeline specifications previously described.^20^ For GenMoz data, raw FASTQ files were downloaded from the Sequence Read Archive and re-run using the same version of the pipeline used for the RegGenE8 data. Sequences resulting from loci targeted by primer pools R1.2 and R2.1 in the *kelch13*, chloroquine resistance transporter (*crt*), multidrug resistance 1 (*mdr1*), dihydropteroate synthase (*dhps*), and dihydrofolate reductase (*dhfr*) genes are reported. Further data cleaning, analysis, and visualization were conducted in R (version 4.3.2). Samples with less than 10,000 reads, targets with <50 reads, and single nucleotide polymorphisms (SNPs) with within-sample-allele-frequency (WSAF) <0.01 or ≤10 reads were excluded from the analysis. Subsequently, SNPs observed in <0.05% of the samples at WSAF <1, and SNPs observed in multiple samples but exclusively in a single sequencing run, were also excluded. Library preparation batches with contaminated negative and/or positive controls were repeated or excluded from all analyses. Samples were considered successfully sequenced if a genotype was obtained for all 59 SNPs evaluated in the *kelch13* gene.

Multilocus haplotypes for polymorphisms in the *dhps* and *dhfr* genes (*dhps*: 436, 437, 540, 581, and 613; *dhfr*: 51, 59, 108, and 164) covered by five independent amplicon targets were identified for each sample by combining all individual microhaplotypes. Haplotypes were identified for samples with either unmixed genotypes at all targets or a single target showing a mixed genotype; in the latter case, the sample was assumed to carry the two haplotypes, accounting for the observed mixture. Samples with mixed genotypes in more than two amplicons were considered indeterminate.

To identify potential issues with sample collection and library preparation, sample similarity was estimated using the root mean square error of WSAF for highly diverse targets (pool D1.1), followed by clustering by Density-Based Spatial Clustering of Applications with Noise (DBSCAN) with ε=0.2. Clusters of samples (at least two samples) with clonality >2 and nearly identical WSAF across all loci, suggesting multiple DBSs were prepared using blood from a single patient, were excluded from the analysis. Multiple such clusters within a RegGenE8 health facility led to the exclusion of all data from that health facility.

#### Statistical Analysis

Descriptive statistics for gender, age group (<5, 5-14, 15-24, 25-39, 40-59, >59 years old), overnight travel in the past 2 months, and country of residence were calculated. Individual participant age was rounded down for age group classification. To enable statistical analysis, some variables were reclassified: farming, herding, and fishing occupations were combined into a single occupational category (agricultural workers); individuals aged 25 years or older were grouped into a single category; and travel reports (within the 2 months prior to sample collection) were categorized as either international or domestic. Differences in socio-demographic characteristics and resistance marker prevalence between countries, as well as between successfully sequenced and qPCR-positive samples, were assessed using Chi-squared tests.

The proportion of samples carrying a mutation or haplotype was calculated as the number of samples with mixed and pure mutant genotypes divided by the total number of genotyped infections.

### Ethical Considerations

Overarching approval for the RegGenE8 study was obtained from the University of California, San Francisco Institutional Review Board (350074), with additional in-country approvals granted by the following authorities: the Angolan Ministry of Health Ethics Committee (34C.E/MINSA.INIS/2022); the Eswatini Health and Human Research Review Board (EHHRRB013/2023); the Namibian Ministry of Health and Social Services Biomedical Research Ethics Committee (22/4/2/3) and the University of Namibia’s Research Ethics Committee (UNAM-DEC-MRS-008-24.08.2022); South Africa’s Human Research Ethics Committees: Medical of the University of the Witwatersrand (M201124), the Mpumalanga Provincial Department of Health (MP_2015RP53_229), the KwaZulu-Natal Provincial Department of Health (KZ_202010_035) and the South Africa National Department of Health; and Zambia’s Tropical Diseases Research Centre Research Ethics Committee (IRB Number 00002911, FWA 3729) and Zambia National Health Research Authority.

The GenMoz project study protocols were approved by the Mozambican National Committee for Bioethics in Health (354/CNBS/2021 and 604/CNBS/21) and Hospital Clínic de Barcelona Ethics Review Committee (HCB/2022/0097).

All samples were collected following informed consent from participants and/or their guardians. No patient consent was required in South Africa as malaria molecular surveillance is conducted as part of routine public health surveillance. All individuals diagnosed with malaria at point of care were treated according to the national treatment guidelines of each participating country. Further details on the consent procedures are available in the country-specific publications.^18–23^

## Results

### Socio-demographic characteristics of the study population

A total of 6,485 samples were used in this regional analysis. In the higher-transmission countries (Angola, Mozambique, and Zambia), the largest proportion of samples (40%) were from participants aged 14 years or younger. In Eswatini and South Africa, both low transmission countries targeting malaria elimination, most study participants were aged 15-39 years (Table 1). Namibia fell between these two groupings, with most study participants aged 5-39 years (Table 1). Gender distribution was balanced in the sample sets from Angola, Mozambique, and Zambia, whereas males predominated in the datasets from the very low transmission countries Eswatini, Namibia, and South Africa (Table 1). Occupational information was rarely recorded in South Africa and was not collected in Mozambique, so they were excluded from the occupation analysis. Although there was a significant difference in occupations among the four remaining countries, (Chi-squared test, p<0.001); 56% of the participants were either minors or students, with most of employed participants working in the agricultural sector (Table 1). International travelers accounted for almost 90% of cases in South Africa and more than one-quarter of cases in Eswatini. In contrast, international travel was uncommon among participants with successfully sequenced samples from Angola, Namibia, and Zambia. Most samples were collected through passive case detection, with 11% collected through active and reactive case detection activities in Eswatini, South Africa, and Zambia.

### Mutations in the kelch13 gene

A total of 15 non-synonymous mutations were detected in the *kelch13* gene, with their prevalence and distribution varying within and between countries. The *kelch13* gene was most diverse in Zambia, where 11 unique mutations were identified. Namibia, despite having the smallest sample size, had nine non-synonymous mutations and the highest proportion of samples carrying a *kelch13* mutation. In contrast, the gene was most conserved in Eswatini, with only a single *kelch13* mutation detected over the study period (Table 2, Figure 2, Table S1 and Figure S1).

**Figure 2.**
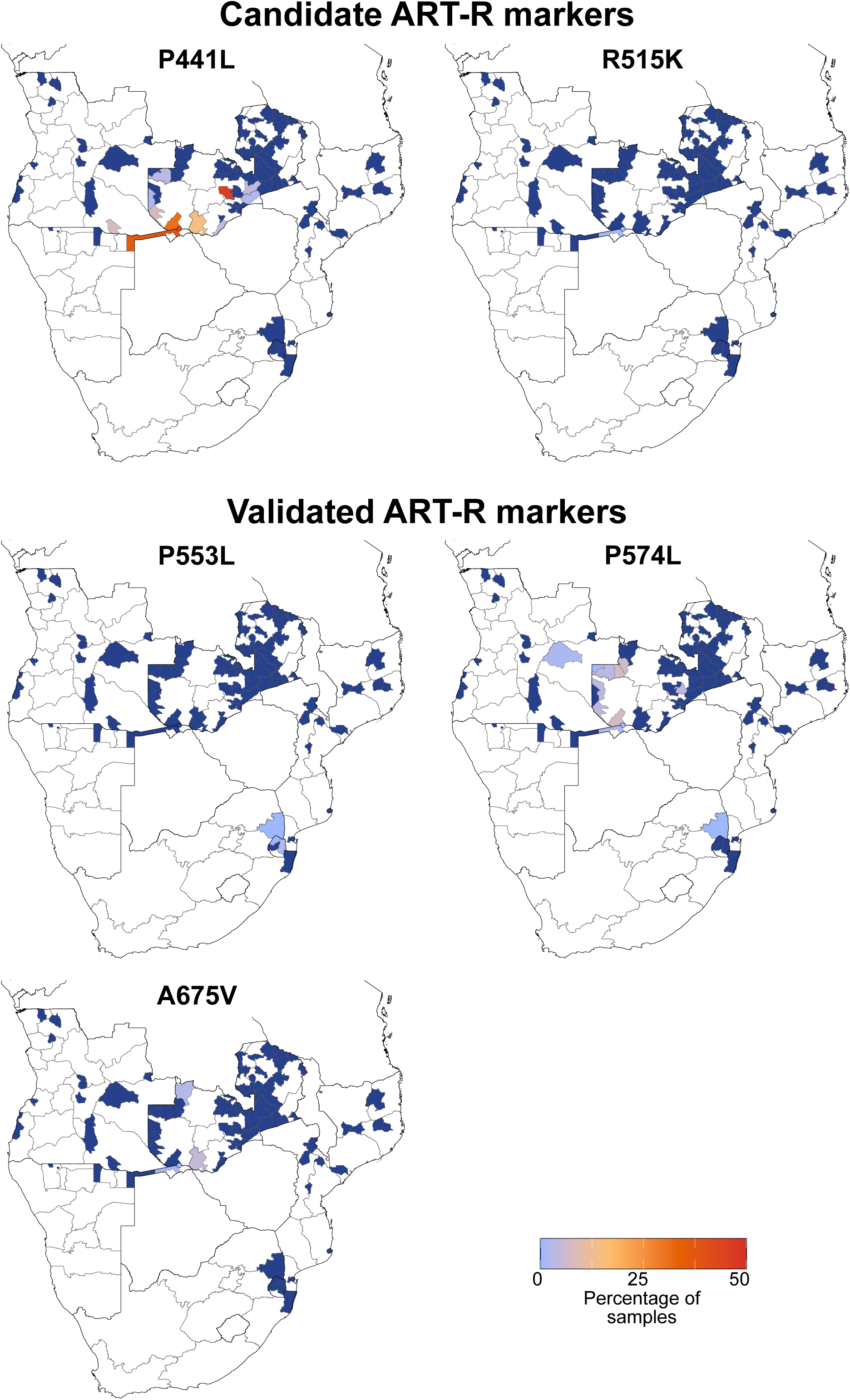
Proportion of samples with validated or candidate artemisinin partial resistance (ART-R) markers in *kelch13* across districts in the six Elimination Eight countries participating in the RegGenE8 and GenMoz studies. Proportions are calculated as the number of samples with a genotype (pure or mixed) divided by the total number of genotyped infections.

**Table 2.** Proportion of samples with non-synonymous mutations in markers of antimalarial drug resistance, across participating Elimination Eight countries. Mutations are shown for artemisinin partial resistance (ART-R; *kelch13*), potential lumefantrine reduced susceptibility (*mdr1*), sulfadoxine-pyrimethamine resistance (*dhfr*, *dhps*), and chloroquine resistance (*crt*, *mdr1*). ART-R markers were categorized according to the current World Health Organization classification framework as validated, candidate, or unvalidated markers of artemisinin partial resistance.^29^ Non-zero values are shown in bold. Proportions are calculated as the number of samples with a genotype (pure or mixed) divided by the total number of genotyped infections.

|  | Angola | Eswatini | Mozambique | Namibia | South Africa | Zambia |
| --- | --- | --- | --- | --- | --- | --- |
| **Validated ART-R markers** |  |  |  |  |  |  |
| *kelch13* P553L | 0%<br>(N=754) | **0.97%**<br>(N=412) | 0%<br>(N=840) | 0%<br>(N=233) | **0.07%**<br>(N=1,522) | 0%<br>(N=2,254) |
| *kelch13* P574L | **0.13%**<br>(N=781) | 0%<br>(N=424) | 0%<br>(N=854) | **1.27%**<br>(N=236) | **0.23%**<br>(N=1,770) | **0.67%**<br>(N=2,400) |
| *kelch13* A675V | 0%<br>(N=711) | 0%<br>(N=382) | 0%<br>(N=847) | **1.3%**<br>(N=231) | 0%<br>(N=1,334) | **0.1%**<br>(N=2,091) |
| **Candidate ART-R markers** |  |  |  |  |  |  |
| *kelch13* R515K | 0%<br>(N=762) | 0%<br>(N=418) | 0%<br>(N=798) | **0.9%**<br>(N=221) | 0%<br>(N=1,589) | 0%<br>(N=2,214) |
| *kelch13* P441L | **0.4%**<br>(N=742) | 0%<br>(N=412) | 0%<br>(N=846) | **32.9%**<br>(N=231) | 0%<br>(N=1,509) | **3%**<br>(N=2,235) |
| **Marker potentially associated with reduced lumefantrine susceptibility** |  |  |  |  |  |  |
| *mdr1* N86 | **99.2%**<br>(N=628) | **100%**<br>(N=336) | **99.88%**<br>(N=836) | **100%**<br>(N=228) | **99.92%**<br>(N=1,207) | **100%**<br>(N=1,949) |
| **SP resistance markers** |  |  |  |  |  |  |
| *dhps* ISGEAA<br>*dhfr* <u>IRNI</u><br>(quintuple) | **25.9%**<br>(N=525) | **93.25%**<br>(N=400) | **91.81%**<br>(N=659) | **72.02%**<br>(N=218) | **97.32%**<br>(N=1,382) | **86.21%**<br>(N=1,827) |
| *dhps* ISGEGA<br>*dhfr* <u>IRNI</u><br>(sextuple) | **1.71%**<br>(N=525) | **6%**<br>(N=400) | **0.61%**<br>(N=659) | **11.01%**<br>(N=218) | **0.29%**<br>(N=1,382) | **3.39%**<br>(N=1,827) |
| **Chloroquine resistance marker** |  |  |  |  |  |  |
| *crt* 72-76 | **18.82%** | **0.47%** | **0.12%** | **0.85%** | **0.06%** | **0.21%** |

| CVIET | (N=781) | (N=425) | (N=862) | (N=235) | (N=1,739) | (N=2,405) |
| --- | --- | --- | --- | --- | --- | --- |

The *kelch13* A578S mutation, not known to be associated with ART-R, was detected across multiple sites and transmission strata in all study countries except Eswatini, but did not exceed 2.5% prevalence in any country. The validated ART-R *kelch13* P574L marker was observed in all countries except Eswatini and Mozambique. This mutation was most prevalent in three high-incidence districts in Zambia’s Western and North-Western provinces (6.4-7.1%). It was also detected in a moderate-incidence district in Zambia’s Central province, as well as in very-low-incidence districts in Namibia’s Zambezi region and Angola’s Moxico province, both bordering Zambia’s Western or North-Western provinces, and in South Africa’s Mpumalanga province. The candidate ART-R marker *kelch13* P441L, detected only in Angola, Namibia, and Zambia, reached proportions exceeding 30% in very-low-incidence settings in Namibia’s Zambezi and Kavango East regions, and Zambia’s Chibombo district (Central province), as well as in the high-incidence Sesheke district in Zambia’s Western province. Outside these districts, *kelch13* P441L was observed at proportions below 15% across settings with transmission ranging from very low to high, primarily along the shared border of Angola, Namibia and Zambia. The mutation was also detected in Zambia’s Lusaka Province (Figure 2).

The uncharacterized *kelch13* P667A mutation had a similar geographical distribution to P441L, reaching proportions above 35% in Zambia’s high-incidence Chavuma and Zambezi districts in North Western Province (Figure 2). Other mutations, including *kelch13* R622T, P667S, and the validated ART-R marker A675V were detected only in Zambia and Namibia. The *kelch13* R622T mutation exceeded 10% in districts with both very-low- and high-incidence in Zambia’s Southern and North-Western provinces. The *kelch13* A675V mutation was detected at low proportions (<5%) in very-low-incidence districts in Zambia’s Southern province and Namibia’s Zambezi region, as well as in a high-incidence district in Zambia’s North-Western province.

The validated ART-R marker *kelch13* P553L was observed in a small cluster of genetically related infections from very-low-incidence settings in Eswatini, including a sample from a traveler from Mozambique and in a single South African sample collected in 2023. This mutation was not observed in Mozambican samples collected in 2022. The candidate ART-R *kelch13* R515K mutation was detected only in Namibia, in less than 1% of samples. Other non-synonymous *kelch13* mutations, including K479I, S485N, V494I, N537S, and Q613E were observed at low proportions across multiple countries and transmission settings.

### Mutations in the dhps and dhfr genes

The proportion of samples carrying the *dhfr* triple mutation (*dhfr* N51I, *dhfr* C59R, and *dhfr* S108N), strongly associated with pyrimethamine resistance, exceeded 85% in all participating countries and was close to fixation in Eswatini, South Africa, and Mozambique (Figure 3, Figure S2, Table S2). However, in districts in southwestern Angola and western Namibia the *dhfr* N51I/C59/S108N haplotype was detected at much lower frequencies, present in just over 20% of samples. In Zambia’s Pemba district, Southern province, three *dhfr* haplotypes, N51I/C59R/S108N; N51I/C59/S108N; and N51/C59R/S108N, were co-circulating at substantial frequencies. The *dhfr* I164L mutation, associated with an approximately 10% increase in pyrimethamine resistance, remains extremely rare in the region and was detected in only two samples from Muchinga province in Zambia.

**Figure 3.**
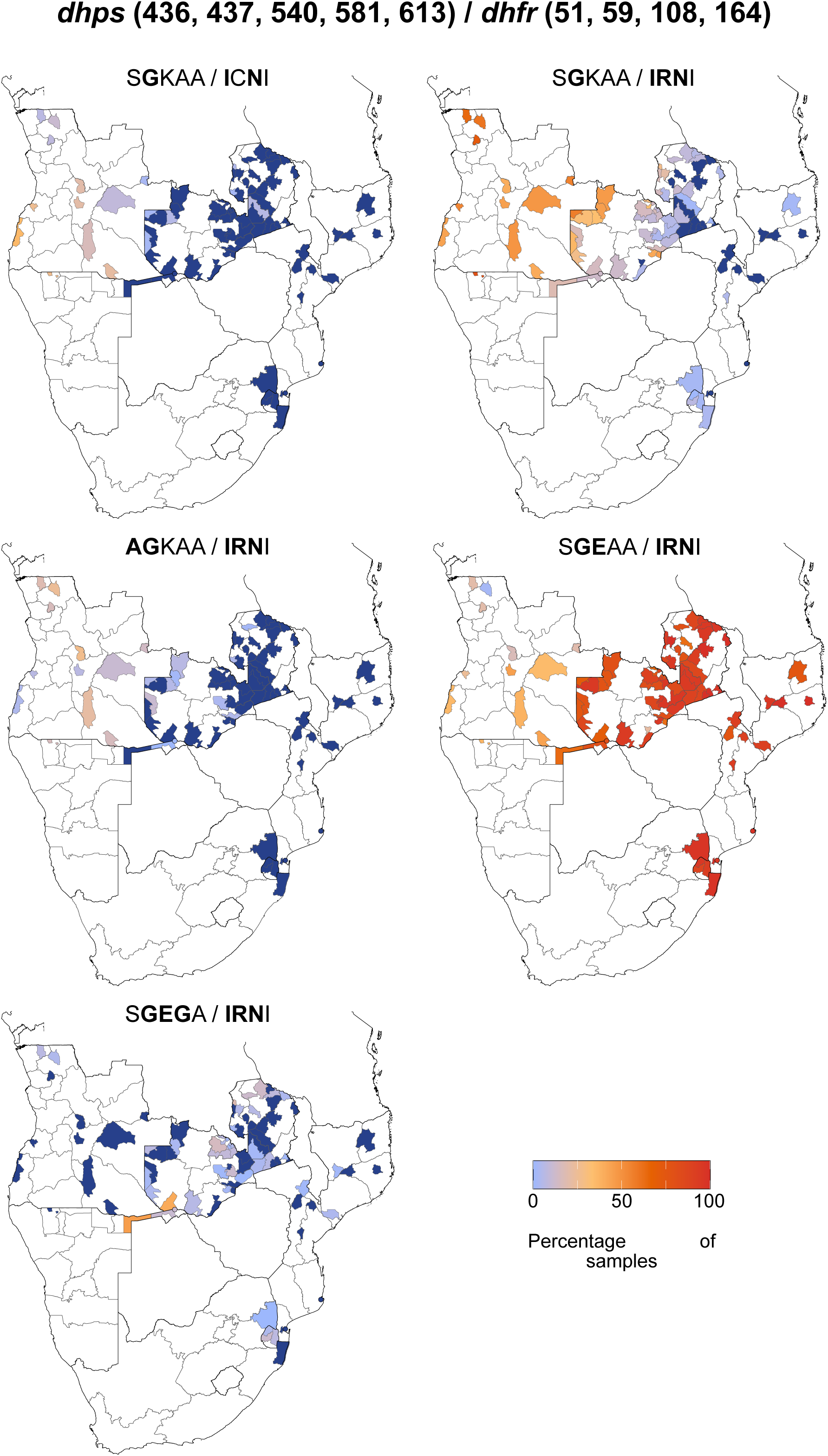
Proportion of samples carrying different *dhps* and *dhfr* combined haplotypes across districts in the six Elimination Eight countries participating in the RegGenE8 and GenMoz studies. Proportions are calculated as the number of samples with a haplotype (pure or mixed) divided by the total number of genotyped infections. Only haplotypes detected in >5% of samples from any country are shown.

In contrast to the *dfhr* triple mutation, the *dhps* double mutation (*dhps* A437G and K540E), associated with sulfadoxine resistance in southern Africa, displayed marked regional heterogeneity.^8^ This mutation ranged from below 40% of samples in Angola to near fixation in South Africa, generally occurring on a haplotype carrying wild-type *dhps* S436, A581, and A613 alleles (Figure 3, Figure S2, Table S2). The second most commonly observed haplotype, *dhps* A437G/K540, was most frequently detected in northwestern Angola, but rarer in the southeastern countries. The *dhps* triple mutation, A437G/K540E/A581G, associated with higher levels of sulfadoxine resistance, was observed in 11.2% of samples from Namibia, 6.1% of samples from Eswatini, and over 10% of samples in several districts in Zambia and in Mbanza-Congo district, Zaire province, northern Angola.^30^ The *dhps* A613S mutation was rare, detected in three samples on the A437G/K540E haplotype, a single sample from Mozambique, South Africa, and Zambia. It was also observed in four samples from Eswatini on a haplotype carrying wild-type *dhps* K540 together with mutant *dhps* I431V, A437G, and A581G alleles.

### Mutations in the crt gene

The distribution of mutations in the *crt* gene was also heterogeneous across the study area (Figure 4, Figure S3, Table S3). The *crt* K76T mutation, associated with chloroquine resistance, was rare in most countries except for Angola, where it was found in 19.1% of samples on a haplotype carrying *crt* M74I and N75E, with wild-type *crt* C72 and V73.^31^ A northwest-to-southeast *crt* K76T mutation gradient was observed, with the mutation most frequent in the northern Angola provinces of Zaire and Uíge (>50% of samples), and decreasing toward the shared borders with Namibia and Zambia (<10%, Figure 4). Similar regional patterns were observed for *crt* A220S, also associated with chloroquine resistance, and *crt* I356T, which is associated with reduced susceptibility to quinine and increased susceptibility to mefloquine (Table S3).^32^ Although generally present at low levels (<3%), the *crt* F48L mutation was detected in all countries except Namibia (Table S2).

**Figure 4.**
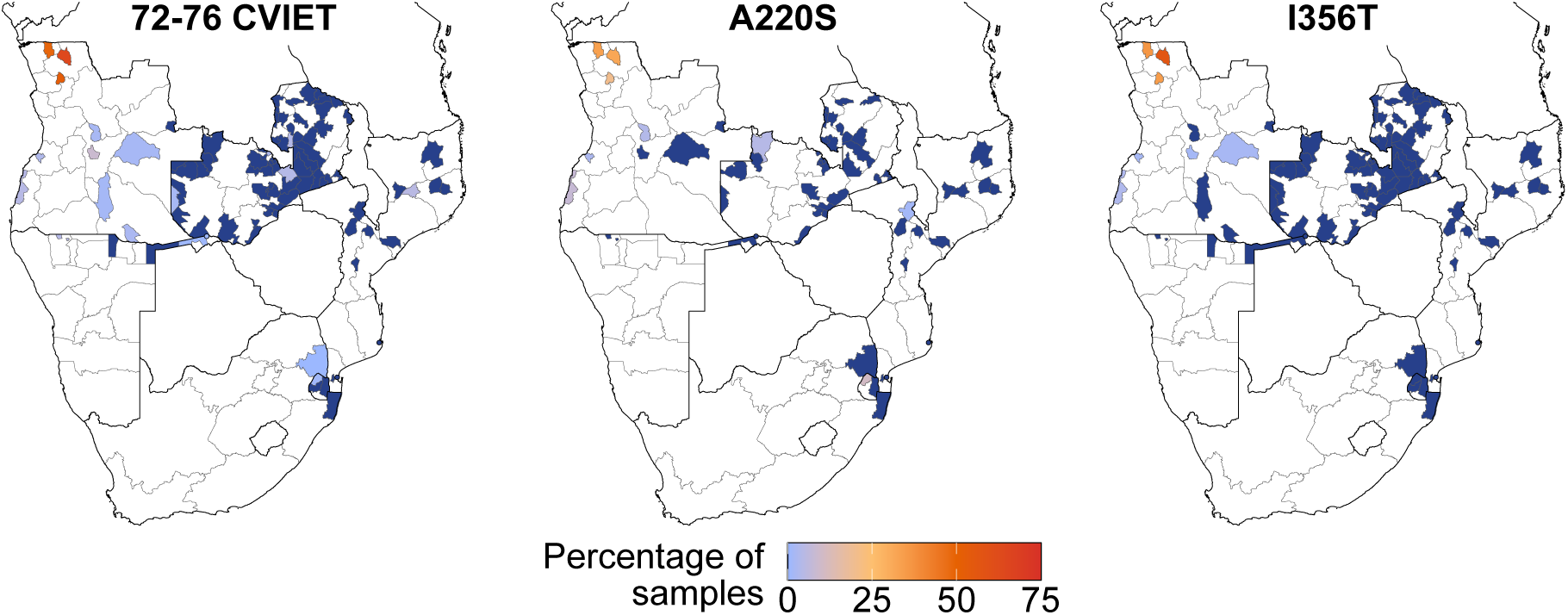
Proportion of samples with mutations in the *crt* gene from the across districts in the six Elimination Eight countries participating in the RegGenE8 and GenMoz studies. Proportions are calculated as the number of samples with a haplotype (pure or mixed) divided by the total number of genotyped infections.

### Mutations in the mdr1 gene

The *mdr1* N86Y mutation, which modulates chloroquine resistance and is associated with susceptibility to lumefantrine, was rarely detected, as was the D1246Y mutation (Table 2, Table S4). In contrast, the *mdr1* Y184F mutation was detected in at least 50% of the samples from each of the participating countries, with the highest frequencies in Eswatini, Mozambique, and South Africa. Among the *mdr*1 haplotypes detected, the wild-type *mdr1* NYD (N86, Y184, and D1246) haplotype was the most prevalent, followed by the single-mutant haplotype NFD (N86, Y184F, and D1246).

Other mutations listed in the WHO compendium of molecular markers for antimalarial drug resistance were either not detected (*crt* C72S, T93S, H97Y, C101F, I218F, N326D, M343L, C350R, G353V, and I356L; *coronin* G50E, R100K, and E107V; *dhps* A16V, *mdr1* S1034C, N1042D) or were not assessed (*crt* G367C, polymorphisms in *cytb* and *mdr1* copy number variations).^33^ The exonuclease gene (*exo*) E415G mutation, associated with piperaquine resistance, was detected in a single sample from Zambia’s Muchinga province.^34^ Polymorphisms hypothesized to constitute a genetic background for ART-R were uncommon, except for the multidrug resistance 2 gene (*mdr2*) I492V mutation, which occurred at district-level proportions ranging from 15-80%, with lower values in southern countries. The *mdr2* T484I mutation was detected in single samples from Zambia and South Africa, while the phosphoinositide-binding protein gene (*pib7*) C1484F and ferredoxin gene (*fd*) D193Y alleles were each detected in one sample from Zambia and South Africa, respectively. The apicoplast ribosomal protein S10 gene (*arps10*) V127M and D128H mutations and *crt* N326S allele were not detected, while the presence of NLI interacting factor-like phosphatase gene (*pph*) V1157L mutation was not evaluated.^35^

## Discussion

By integrating molecular surveillance data from six southern African countries generated through harmonized sampling and analytical approaches, this analysis provides the first regional synthesis of antimalarial drug resistance marker distribution across southern Africa. The findings have direct implications for case management and chemoprevention policies, and identify priority areas for research and therapeutic efficacy studies.

Although validated *kelch13* ART-R markers remained rare across the study region, never exceeding 1.3% in any country, pronounced spatial heterogeneity at the sub-national level was detected. Validated and candidate ART-R markers, as well as uncharacterized *kelch13* mutations, were concentrated in districts along the shared Angola-Namibia-Zambia border and in central Zambia. Notably, the candidate ART-R marker *kelch13* P441L exceeded 30% in districts in eastern Namibia and Zambia’s Central and Western provinces. This mutation has now been reported across multiple eastern African countries with evidence of positive selection, though its clinical relevance in an African setting requires further investigation.^36–38^ It is also unclear whether the southern African lineages share a common ancestor with eastern African lineages or arose independently.

Outside the Angola-Namibia-Zambia hotspot, validated ART-R markers in the other study countries were rare. A small cluster of genetically related *kelch13* P553L infections from Eswatini and a single case from South Africa, with travel histories suggesting importation from Mozambique was detected. However, this mutation was not detected in Mozambican samples from the preceding transmission season. As this mutation has not been detected in South Africa since, it is possible that this mutation alone offered no selective advantage in southern Africa.

The epidemiological, ecological, and biological factors underpinning ART-R emergence and spread are not yet fully understood. Drug pressure in low-transmission settings, where host immunity is lower and parasite competition is reduced, has been hypothesized to facilitate the emergence of resistance.^39^ However, in this regional analysis, *kelch13* mutations were observed across all transmission strata. The Angola-Namibia-Zambia hotspot is characterized by highly heterogeneous and seasonal transmission intensities, with very-low transmission districts, prone to malaria outbreaks bordering high-transmission areas.

Southern Africa, like all regions in Africa, is characterized by highly mobile populations, whose mobility patterns shape malaria transmission and connectivity across the region.^40^ These vulnerable populations often face barriers to accessing the appropriate care and malaria prevention tools, which, in conjunction with occupational and/or behavioral exposure facilitates the movement of parasites between areas.^41,42^ Such movement may have contributed to the observed distribution of *kelch13* mutations in Namibia and Zambia. However, other southern African regions with similar low transmission intensities and population mobility, including eliminating districts in Eswatini, Mozambique, and South Africa, showed only limited evidence of validated ART-R markers. This suggests that drug pressure and malaria importation alone are insufficient to explain the *kelch13* P441L hotspots detected in this analysis. Differences in parasite genetic background, access to care, antimalarial drug stewardship, and other contextual factors likely also contribute. Deep sequencing of archived samples across multiple transmission seasons, integrated with human mobility and transmission intensity data, would enable reconstruction of the evolutionary history of these mutations in this region.

The wildtype *mdr1* N86 allele was at or nearing fixation across all six countries. The clinical impact of this allele on lumefantrine efficacy remains unresolved. A meta-analysis showed an association with artemether-lumefantrine treatment failures, while in vitro investigations revealed only modest changes in lumefantrine susceptibility.^43,44^ A recently described haplotype in the *px1* gene associated with decreased lumefantrine susceptibility was not evaluated here.^45^ No mutations associated with resistance to piperaquine or amodiaquine, other ACT partner drugs, were detected. Copy number variation markers associated with piperaquine and lumefantrine resistance were not assessed.^33^

Encouragingly, efficacies above 90% have been reported for artemether-lumefantrine, the preferred ACT in all six study countries, across Mozambique, and in Zambia (NMEC, personal communication).^46^ However, in three provinces in Angola (Bengo, Lunda Sul and Zaire), efficacies below the WHO 90% threshold have been observed.^47,48^ Interestingly, no validated or candidate ART-R markers were detected in Zaire province in this or prior studies reporting reduced efficacy, suggesting treatment failures in these areas are not driven primarily by ART-R. The recent observation of *mdr1* gene duplications in Angolan treatment failures points toward reduced lumefantrine susceptibility as a potential contributor.^48^

In the districts where *kelch13* mutations were identified, parasites predominantly carried the *mdr1* N86 wildtype allele potentially associated with reduced lumefantrine susceptibility. Although clinical failures have not yet been widely reported in these areas, the combination of emerging *kelch13* variants and a potentially lumefantrine-tolerant genetic background warrants close monitoring. Erosion of partner drug efficacy may precede or potentiate ACT resistance.^49^ However, the low prevalence of ART-R markers combined with high artemether-lumefantrine efficacy data, suggests that there is currently no urgent concern for ACT efficacy across the six participating countries outside the localized areas reporting reduced artemether-lumefantrine efficacy or elevated *kelch13* mutation frequencies.

While SP resistance does not directly threaten first line ACT efficacy, its regional distribution reinforces the broader picture of a resistance landscape shaped by drug pressure gradients and cross-border parasite movement. The prevalence of the *dhfr*-*dhps* quintuple mutation was markedly heterogeneous across the region, with Angola and northwestern Namibia showing substantially lower SP resistance prevalence than the southeastern countries, where it approached fixation. This pattern positions Angola and Namibia at the transition zone within the continental west-to-east gradient of low to high *dhps* K540E prevalence.^50^ The quintuple haplotype combined with the *dhps* A581G mutation, associated with reduced SP-intermittent preventive treatment (IPT) in pregnancy efficacy, exceeded 5% in Eswatini, Namibia, Angola (Zaire province), and several Zambian districts.^51,52^ As SP is not used for treatment or chemoprevention in Eswatini and Namibia it is unclear what is driving the selection of the *dhps* A581G mutation in these countries. National malaria programs implementing SP-IPT in pregnancy should consider chemopreventive efficacy studies and potential regimen changes in areas where this sextuple haplotype is prevalent.

Several study limitations warrant consideration. Sampling for most countries covered a single transmission season, limiting the ability to assess temporal trends in resistance marker selection at a regional level. The mismatch in GenMoz and RegGenE8 sampling periods (Mozambique in 2022 versus most other countries in 2023) may have obscured cross-border resistance signals; contemporaneous sampling across all participating countries should be a priority for future regional studies. Both Botswana and Zimbabwe declined to participate in RegGenE8, creating a corridor of uncertainty between the Angola-Namibia-Zambia block and the Eswatini-Mozambique-South Africa block. Whether *kelch13* mutations have spread into these countries, and how far south they may be present, remains unknown. To the best of our knowledge, no recent TES data are available from Zimbabwe, further limiting the ability to assess ACT efficacy across this corridor. Generating resistance and efficacy data for both countries is an urgent priority. Finally, the studies assessed only validated markers of resistance; investigation of treatment failures through genomic analysis of recrudescent parasites remains the most direct path to identifying and characterizing novel markers with clinical relevance.^53^

The molecular landscape described here provides valuable insight into antimalarial drug resistance marker distribution across six southern African countries that can be used to guide preparedness planning for resistance prevention and containment. In districts where *kelch13* mutation frequencies are rising, ensuring effective treatment availability is of paramount importance. Modeling studies suggest that multiple first-line therapy strategies and triple ACTs are among the most effective single policies for slowing the spread of ART-R and reducing treatment failures.^54,55^ Rapid data sharing between neighboring national malaria programs and the generation of maps that integrate parasite genomic and human mobility data with transmission intensity will support with the identification of districts most at risk, enabling prospective targeting of preventative interventions. As the region moves toward elimination, sustaining and expanding the molecular surveillance infrastructure established by RegGenE8 and GenMoz to include all E8 countries, will be essential to protect treatment efficacy and support elimination goals across southern Africa.

## Data Availability

Primary data were sourced from previously published studies cited herein

## Acknowledgements

The authors thank the participants, nurses, field supervisors and data managers that made this work possible. We also thank Nokwethemba Kubheka and Ongeziwe Taku from the National Institute for Communicable Diseases for their technical support. We also thank members of the Malaria Elimination Initiative, the Experimental and Population-based Pathogen Investigation Center, and the Infectious Disease Dynamics group at Johns Hopkins University for productive discussions. Finally, we are grateful to the Southern African Development Community Malaria Elimination Eight for advocacy and coordination of this work across the region.

## Financial support

This work was supported by the Bill & Melinda Gates Foundation (INV-024346, INV-019032, INV-067310, INV 031512), the Global Fund (QPA-M-E8S), the Departament d’Universitats i Recerca de la Generalitat de Catalunya (AGAUR; grant 2021 SGR 01517, and grant 2022 FI_B 00148). JLS was supported by an award from the National Institutes of Health/National Institute of Allergy and Infectious Diseases grant number 5K01AI153555. The Centro de Investigaçâo em Saude de Manhica (CISM) is supported by the Government of Mozambique and the Spanish Agency for International Development Cooperation (AECID). This research is part of ISGlobal’s Program on the Molecular Mechanisms of Malaria which is partially supported by the Fundación Ramón Areces. We acknowledge support from the grant CEX2023-0001290-S funded by MCIN/AEI/10.13039/501100011033, and support from the Government of Catalonia through the CERCA Program.

## Conflict of Interest

The authors declare no conflict of interest.

## Co-author Contact Information

## Notes

### Competing Interest Statement

The authors have declared no competing interest.

### Author Declarations

Ethics committee/IRB of University of California, San Francisco gave ethical approval for this work (IRB 350074). Ethics committee/IRB of Angolan Ministry of Health gave ethical approval for this work (34C.E/MINSA.INIS/2022). Ethics committee/IRB of Eswatini Health and Human Research Review Board gave ethical approval for this work (EHHRRB013/2023). Ethics committee/IRB of Namibian Ministry of Health and Social Services gave ethical approval for this work (22/4/2/3). Ethics committee/IRB of University of Namibia gave ethical approval for this work (UNAM-DEC-MRS-008-24.08.2022). Ethics committee/IRB of University of the Witwatersrand gave ethical approval for this work (M201124). Ethics committee/IRB of Mpumalanga Provincial Department of Health gave ethical approval for this work (MP_2015RP53_229). Ethics committee/IRB of KwaZulu-Natal Provincial Department of Health gave ethical approval for this work (KZ_202010_035). Ethics committee/IRB of South Africa National Department of Health gave ethical approval for this work. Ethics committee/IRB of Tropical Diseases Research Centre gave ethical approval for this work (IRB 00002911, FWA 3729). Ethics committee/IRB of Zambia National Health Research Authority gave ethical approval for this work. Ethics committee/IRB of Mozambican National Committee for Bioethics in Health gave ethical approval for this work (354/CNBS/2021 and 604/CNBS/21). Ethics committee/IRB of Hospital Clinic de Barcelona gave ethical approval for this work (HCB/2022/0097).

