## Supplementary Material for "Geographical Variation in Antimalarial Drug Resistance Marker Prevalence Across the Southern African Elimination Eight Region"

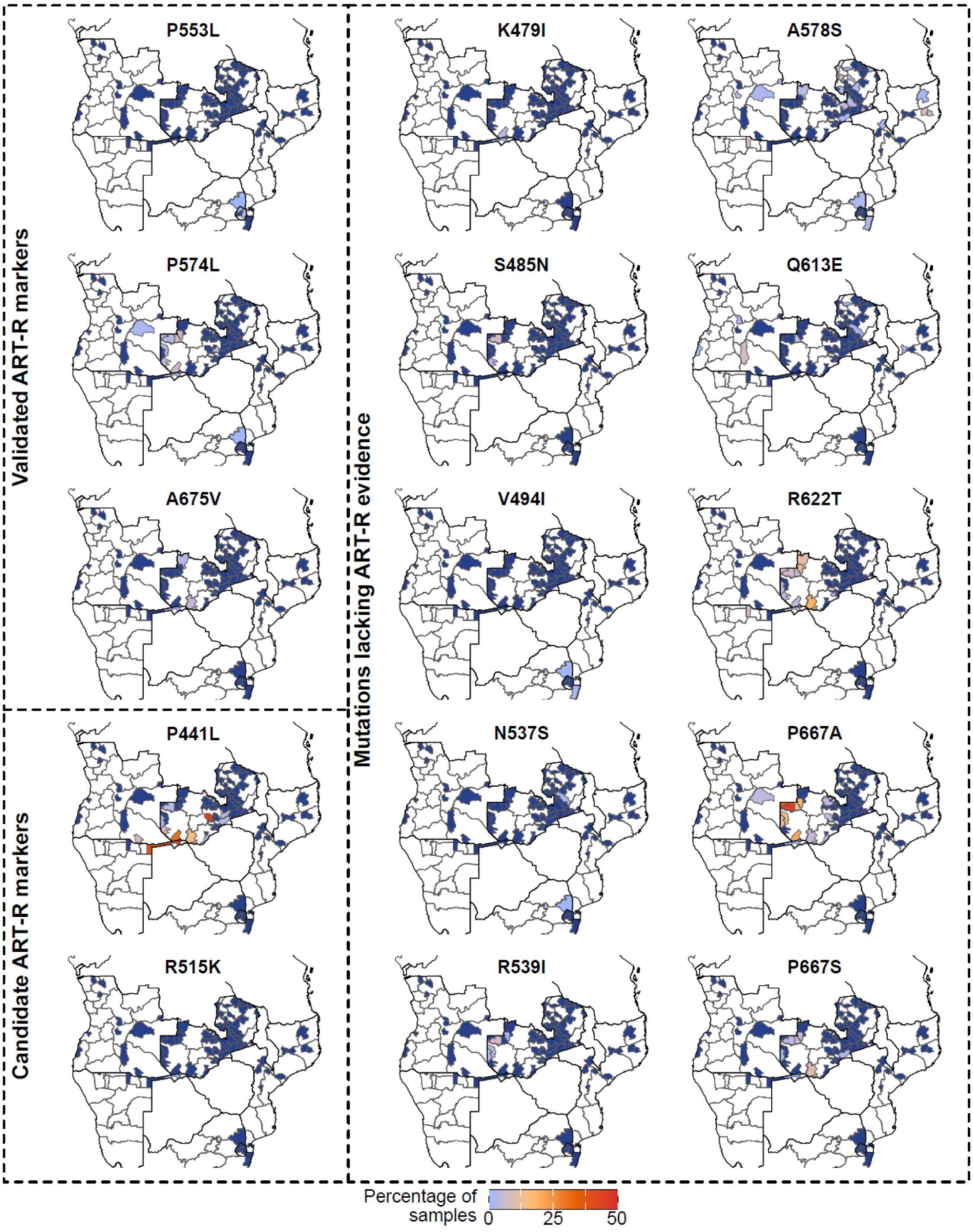


**Figure S1. Proportion of samples with non-synonymous *kelch13* mutations in the study districts from the six participating Elimination Eight countries.** Proportions are calculated as the number of samples with a genotype (pure or mixed) divided by the total number of genotyped infections.

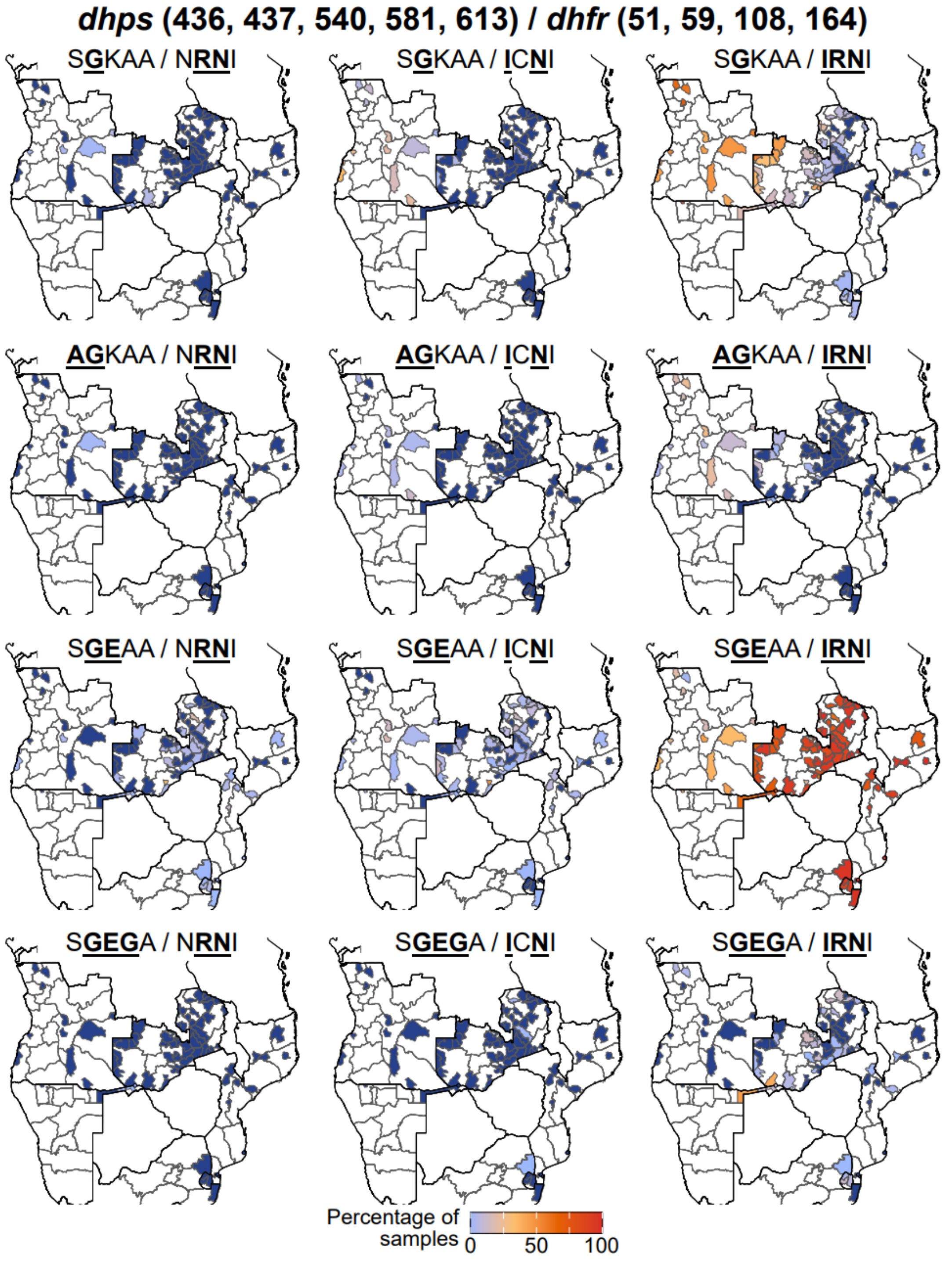


**Figure S2. Proportion of samples from the study districts in the six participating Elimination Eight countries carrying different *dhps* and *dhfr* combined haplotypes.**

Proportions are calculated as the number of samples with a haplotype (pure or mixed) divided by the total number of genotyped infections.

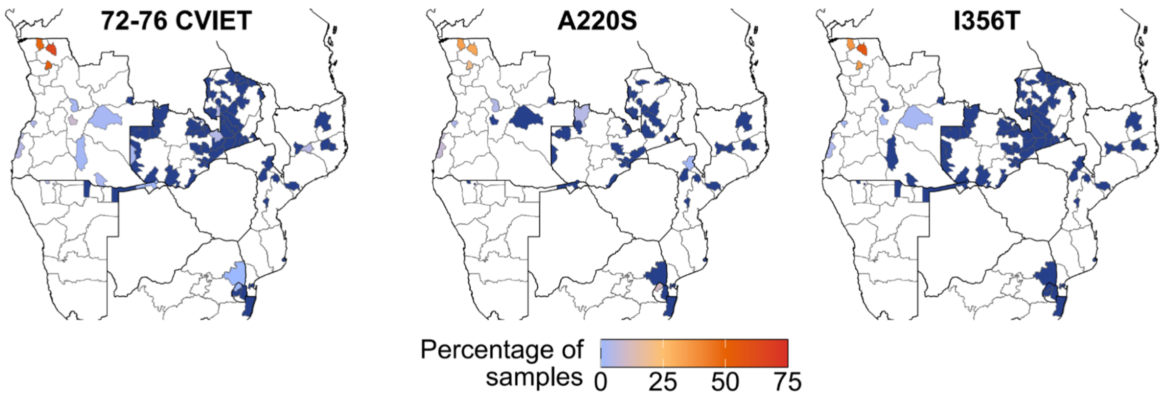


**Figure S3. Proportion of samples from the study districts in the six participating Elimination Eight countries with mutations in the *crt* gene.**

**Table S1. Proportion of samples with non-synonymous *kelch13* mutations by mutation and participating Elimination 8 countries.** Mutations were categorized using the current World Health Organization classification framework as validated, candidate, or unvalidated markers of artemisinin partial resistance (ART-R).[29] Non-zero values are shown in bold. Proportions are calculated as the number of samples with a genotype (pure or mixed) divided by the total number of genotyped infections.

| | Angola | Eswatini | Mozambique | Namibia | South Africa | Zambia |
|---|---|---|---|---|---|---|
| **Validated ART-R markers** | | | | | | |
| **P553L** | 0% (N=754) | **0.97% (N=412)** | 0% (N=840) | 0% (N=233) | **0.07% (N=1,522)** | 0% (N=2,254) |
| **P574L** | **0.13% (N=781)** | 0% (N=424) | 0% (N=854) | **1.27% (N=236)** | **0.23% (N=1,770)** | **0.67% (N=2,400)** |
| **A675V** | 0% (N=711) | 0% (N=382) | 0% (N=847) | **1.3% (N=231)** | 0% (N=1,334) | **0.1% (N=2,091)** |
| **Candidate ART-R markers** | | | | | | |
| **R515K** | 0% (N=762) | 0% (N=418) | 0% (N=798) | **0.9% (N=221)** | 0% (N=1,589) | 0% (N=2,214) |
| **P441L** | **0.4% (N=742)** | 0% (N=412) | 0% (N=846) | **32.9% (N=231)** | 0% (N=1,509) | **3% (N=2,235)** |
| **Mutations lacking evidence of ART-R** | | | | | | |
| **K479I** | 0% (N=781) | 0% (N=424) | 0% (N=853) | **0.86% (N=233)** | 0% (0/1,719) | **0.04% (N=2,389)** |
| **S485N** | 0% (N=781) | 0% (N=424) | 0% (N=853) | 0% (N=233) | 0% (N=1,719) | **0.21% (N=2,389)** |
| **V494I** | 0% (N=781) | 0% (N=424) | **0.12% (N=853)** | 0% (N=233) | **1.11% (N=1,719)** | 0% (N=2,389) |
| **N537S** | 0% (N=754) | 0% (N=412) | 0% (N=840) | 0% (N=233) | **0.39% (N=1,522)** | **0.04% (N=2,254)** |
| **R539I** | 0% (N=754) | 0% (N=412) | 0% (N=840) | 0% (N=233) | 0% (N=1,522) | **0.22% (N=2,254)** |
| **A578S** | **0.77% (N=781)** | 0% (N=424) | **0.7% (N=854)** | **0.42% (N=236)** | **2.37% (N=1,770)** | **0.92% (N=2,400)** |
| **Q613E** | **1.28% (N=781)** | 0% (N=422) | **0.12% (N=823)** | 0% (N=236) | 0% (N=1,718) | **0.17% (N=2,331)** |
| **R622T** | 0% (N=711) | 0% (N=382) | 0% (N=847) | **4.33% (N=231)** | 0% (N=1,334) | **1.48% (N=2,091)** |
| **P667A** | **0.28% (N=711)** | 0% (N=382) | 0% (N=847) | **0.87% (N=231)** | 0% (N=1,334) | **3.4% (N=2,091)** |
| **P667S** | 0% (N=711) | 0% (N=382) | 0% (N=847) | **0.43% (N=231)** | 0% (N=1,334) | **0.53% (N=2,091)** |

**Table S2. Proportions of samples from each of the participating Elimination Eight countries with mutations in the *dhps* and *dhfr* genes.** Proportions are calculated as the number of samples with a genotype (pure or mixed) divided by the total number of genotyped infections. Undetermined haplotypes, and haplotypes present in <0.5% of the samples in any country are excluded. Nonsynonymous mutations are underlined and in bold

| | Angola | Eswatini | Mozambique | Namibia | South Africa | Zambia |
|---|---|---|---|---|---|---|
| ***dhfr* (51, 59, 108 ,164)** | N=748 | N=421 | N=840 | N=232 | N=1593 | N=2282 |
| N**RN**I | **2.94%** | **3.56%** | **3.93%** | **5.17%** | **1.32%** | **3.77%** |
| **I**C**N**I | **36.9%** | 0% | **3.1%** | **8.19%** | **1%** | **7.1%** |
| **IRN**I | **87.17%** | **99.05%** | **98.33%** | **96.12%** | **99.56%** | **96.93%** |
| ***dhps* (431, 436, 437, 540, 581,613)** | N=622 | N=400 | N=668 | N=224 | N=1391 | N=1869 |
| ISAKAA | **3.38%** | **2.5%** | **4.79%** | 0% | **1.65%** | **2.62%** |
| I**A**AKAA | **0.48%** | 0% | **1.05%** | 0% | **0.14%** | **0.27%** |
| IS**G**KAA | **69.61%** | 2% | **0.75%** | **25.45%** | **2.95%** | **17.17%** |
| I**AG**KAA | **21.38%** | 0% | 0% | **2.23%** | 0% | **0.91%** |
| I**AGE**AA | **2.09%** | 0% | **0.15%** | 0% | 0% | **0.05%** |
| ISA**E**AA | 0% | **0.75%** | **0.75%** | 0% | **0.86%** | **0.7%** |
| IS**GE**AA | **32.48%** | **94.25%** | **93.41%** | **74.11%** | **97.63%** | **89.73%** |
| IS**GEG**A | **1.29%** | **6%** | **0.75%** | **11.16%** | **0.36%** | **3.42%** |
| **V**A**G**K**GS** | 0% | **0.75%** | 0% | 0% | 0% | 0% |
| ***dhps dhfr*** | N=525 | N=400 | N=659 | N=218 | N=1382 | N=1827 |
| ISAKAA **I**C**N**I | **0.76%** | 0% | 0% | 0% | **0.07%** | **0.22%** |
| ISAKAA **IRN**I | **1.9%** | **2.5%** | **4.55%** | 0% | **1.59%** | **2.35%** |
| I**A**AKAA **IRN**I | **0.38%** | 0% | **0.91%** | 0% | **0.14%** | **0.22%** |
| IS**G**KAA N**RN**I | **1.14%** | 0% | 0% | **0.92%** | 0% | **0.05%** |
| IS**G**KAA **I**C**N**I | **16.38%** | 0% | 0% | **4.59%** | 0% | **0.33%** |
| IS**G**KAA **IRN**I | **53.52%** | **2%** | **0.61%** | **21.56%** | **2.68%** | **15.27%** |
| I**AG**KAA **I**C**N**I | **3.81%** | 0% | 0% | 0% | 0% | **0.05%** |
| I**AG**KAA **IRN**I | **14.29%** | 0% | 0% | **2.29%** | 0% | **0.93%** |
| I**AGE**AA **IRN**I | **1.52%** | 0% | 0% | 0% | 0% | **0.05%** |
| ISA**E**AA **IRN**I | 0% | **0.75%** | **0.61%** | 0% | **0.87%** | **0.6%** |
| IS**GE**AA N**RN**I | **0.38%** | **3.5%** | **2.88%** | **1.83%** | **0.72%** | **2.63%** |
| IS**GE**AA **I**C**N**I | **3.43%** | 0% | **2.28%** | **2.29%** | **0.65%** | **4.27%** |
| IS**GE**AA **IRN**I | **25.9%** | **93.25%** | **91.81%** | **72.02%** | **97.32%** | **86.21%** |
| IS**GEG**A **IRN**I | **1.71%** | **6%** | **0.61%** | **11.01%** | **0.29%** | **3.39%** |
| **V**A**G**K**GS** **IRN**I | 0% | **0.75%** | 0% | 0% | 0% | 0% |

**Table S3. Proportions of samples from each of the participating Elimination Eight countries with mutations in the *crt* gene.**

Proportions are calculated as the number of samples with a genotype (pure or mixed) divided by the total number of genotyped infections. Only mutations/haplotypes present in >0.5% of the samples in any country are shown

| | **Angola** | **Eswatini** | **Mozambique** | **Namibia** | **South Africa** | **Zambia** |
|---|---|---|---|---|---|---|
| 72-76 CVIET | 18.82% (N=781) | 0.47% (N=425) | 0.12% (N=862) | 0.85% (N=235) | 0.06% (N=1,739) | 0.21% (N=2,405) |
| A220S | 12.56% (N=223) | 5.26% (N=95) | 0.14% (N=732) | 0% (N=130) | 0% (N=545) | 0.13% (N=747) |
| I356T | 13.69% (N=745) | 0% (N=423) | 0% (N=854) | 0% (N=228) | 0% (N=1,564) | 0% (N=2,300) |

**Table S4. Proportions of samples from each of the participating Elimination Eight countries with mutations in the *mdr1* gene**

Proportions are calculated as the number of samples with a genotype (pure or mixed) divided by the total number of genotyped infections.

| | Angola | Eswatini | Mozambique | Namibia | South Africa | Zambia |
|---|---|---|---|---|---|---|
| **86** | N=628 | N=336 | N=836 | N=228 | N=1,207 | N=1,949 |
| N | 99.2% | 100% | 99.88% | 100% | 99.92% | 100% |
| Y | 2.07% | 0% | 0.12% | 0% | 0.99% | 0.21% |
| **184** | N=760 | N=421 | N=854 | N=232 | N=1,460 | N=2,273 |
| F | 52.11% | 66.27% | 72.13% | 51.72% | 67.47% | 52.27% |
| **1246** | N=628 | N=336 | N=836 | N=228 | N=1,207 | N=1,949 |
| Y | 2.07% | 0% | 0.12% | 0% | 0.99% | 0.21% |
| **86, 184, 1246 haplotype** | N=621 | N=335 | N=827 | N=224 | N=1,164 | N=1,937 |
| NFD | 51.05% | 65.97% | 71.95% | 50.89% | 67.7% | 52.35% |
| NYD | 79.07% | 58.51% | 70.25% | 70.98% | 69.85% | 81.52% |
| YYD | 1.13% | 0% | 0% | 0% | 0.09% | 0% |
